# Association of polycyclic aromatic hydrocarbons in moss with blood biomarker among nearby residents in Portland, Oregon

**DOI:** 10.1101/2021.07.15.21260592

**Authors:** Igor Burstyn, Geoffrey H. Donovan, Yvonne L. Michael, Sarah Jovan

## Abstract

Polycyclic aromatic hydrocarbons (PAHs) are a component of air pollutants that are costly to measure using traditional air-quality monitoring methods. We used an epiphytic bio-indicator (moss genus: *Orthotrichum*) to cost-effectively evaluate atmospheric deposition of PAHs in Portland, Oregon in May 2013. However, it is unclear if measurements derived from these bioindicators are good proxies for human exposure. To address this question, we simultaneously, measured PAH-DNA adducts in blood samples of non-smokers residing close to the sites of moss measurements. We accounted for individual determinants of PAH uptake that are not related to environmental air quality through questionnaires, e.g., wood fires, consumption of barbecued and fried meats. Correlation and linear regression (to control for confounders from the lifestyle factors) evaluated the associations. We did not observe evidence of an association between PAH levels in moss and PAH-DNA adducts in blood of nearby residents, but higher level of adduct were evident in those who used wood fire in their houses in the last 48 hours. It remains to be determined whether bio-indicators in moss can be used for human health risk assessment.

**Highlights:**

- Concentration of PAH in moss do not relate to DNA adducts of PAH in blood of nearby residents in our sample.
- Content of moss may not be suitable for assessing exposure to PAH among humans.

## Introduction

Polycyclic aromatic hydrocarbons (PAHs) are a class of air pollutants, some of which are linked to adverse health outcomes (1, 2). PAHs are by-products of the incomplete combustion of organic matter, including tobacco products, fossil fuels and firewood, and are abundant in smoked, fried, or grilled food. Risk management requires understanding pathways of exposure, best done though comparing environmental measurements that reflect different sources to measurements of internal dose through biomarkers. Cost-effective methods to accurately evaluate exposures to PAH are lacking, unlike that for total particle matter, typically employed in air pollution research and regulations.

Lichen and moss are among the most commonly used bio-indicators of atmospheric PAHs and can accumulate both particle and gas-phase PAHs (3). Their leaves and lobes lack cuticles, allowing PAHs to diffuse easily into cells (4) and a high-surface area traps particles (5). In addition, lichen and moss do not have roots, making them dependent on atmospheric sources of nutrients and water and, therefore, bioaccumulation reflects airborne sources of PAHs.

Lichens and moss contain levels of mid-to-high molecular weight PAHs that correlate with nearby stationary air-quality monitors in studies conducted by different groups across diverse locations (3, 5, 6). Jovan et al. (7) demonstrated that moss can be used to cost-effectively map distribution of PAHs across Portland, Oregon; earlier related research on metals indicated the same moss species, *Orthotrichum lyellii* (Hook & Taylor), can be used to identify sources of such pollution (8). It is uncertain how PAH levels in lichens and moss relate to human exposures. To increase our understanding of this critical question, we conducted a follow-up study to Jovan et al. (7) investigating the relationship between levels of PAHs in moss and that of PAH-DNA adducts in the blood of nearby residents.

## Materials and Methods

### Study area and sampling strategy

Portland is a city in northwest Oregon, USA; *Orthotrichum lyellii* is the only species of moss or lichen that is abundant across a wide variety of sites in Portland. Participants were non-smokers residing with non-smokers recruited though an email list distributed to employees of US Forest Service’s Pacific Northwest Research Station located in Portland. We collected moss samples in May of 2013 on hardwood trees located on properties of study participants, from a height of at least 1 meter to reduce the influence of spray from cars and from dog urine. We recorded the location of each sample point using a high-accuracy GPS (Garmin Oregon 450). Drexel University Institutional Review Board approved the study protocol.

### Human biological monitoring and questionnaires

Participants completed questionnaires that capture demographics and exposure to known sources of PAH during the preceding 48 hours: wood fires in house, consumption of fried, barbecued, grilled, or smoked food.

A licensed phlebotomist collected 50 ml of blood by venipuncture. Blood samples were transported by overnighted delivery for quantification of PAH-DNA adduct at the laboratory at Columbia University’s Mailman School of Public Health following protocol of Santella et al. (9-11).

### Moss sample preparation and testing

Methods of moss sampling and testing are reported in detail by Jovan et al. (7) and are only briefly summarized here. Immediately after collection, we stored samples at 4°C in metalized polyester Kapak bags. On a sterilized lab bench, we removed debris and trimmed off most brown tissue. On average, samples were stored for one week or less before analysis.

We ground samples in the lab, using an ASE 200 (Dionex, Sunnyvale, CA) for pressurized liquid extraction of PAHs. We loaded moss samples into stainless steel cells, with 1 gram of Florisil® (activated magnesium silicate; U.S. Silica Company, Berkeley Springs, USA) placed at the outlet of the cell. We used dichloromethane for PAH extraction. We pre-heated cells to 100^0^C for five minutes then processed with two static extraction cycles. Between cycles, we flushed the cells with 50% of cell volume solvent. At the end of cycles, the cell was flushed with high purity nitrogen (N). We concentrated the PAH extract under N at ambient temperature to a final volume of 1mL.

We analyzed the extract using a method similar to EPA 8270D, utilizing an Agilent 6890GC with 5973 Mass Selective Detector, operating in full scan mode, and a Rested Rxi-XLB column. For quality assurance, we ran a method blanks, and spiked each sample with isotopically labeled (deuterated) PAH surrogates that served as internal standards to monitor calibration and performance of the GC-MS instrument.

### Statistical Analysis

We replaced all non-detectable samples values with half of the limit of detection. We estimated Spearman rank correlation of measurements of PAH in moss and blood adducts that were detected. We explored the correlation structure among PAH levels in moss via principal components analysis (PCA) of detected levels of PAH and used it to limit the number of PAH in moss that need to be considered in relation to PAH-DNA adducts. The number of principal components that we considered in analysis was indicated by examination of the scree plot. PAHs representative of each selected principal component i.e., those with the highest loadings, were used as predictors of blood PAH-DNA adduct levels. Because PAH-DNA were right-skewed (appeared to be log-normal), we applied logarithmic transformation to ensure validity of parametric statistical tests that require assumption of Gaussian distribution. Associations of PAH-DNA adducts in the blood were related to PAH levels in moss using linear regression controlling for wood burning in the house and consumption of smoked, grilled, or barbecued foods, as well as race (white vs. other), age (continuous in years) and gender; all covariates were selected *a priori*.

## Results

We recruited 53 volunteers for the study, all of whom were able to provide blood samples and had trees on their properties that yielded moss samples. The volunteers were predominantly female (n=31), white (n=47) and aged on average 51 years (range 30 to 68). Only 3 reported wood fires on their properties and 17 reported eating fried or grilled food within 48 hours of the measurements.

In quality control experiments, average percent recoveries of known concentrations of isotopically labeled surrogate PAH compounds added to moss samples were all indistinguishable from 100%, within one standard deviation; all method blanks were at or below detection (details not shown). The level of PAH-DNA adducts and PAH levels in moss are summarized in **Table 1**. Among the 32 measurements of adducts that were above the detection limit (5.6 adducts/10^8^nucleotides), the average level was 11.8 adducts/10^8^nucleotide with a range of 7.9 to 22.9; coefficient of variation (CV) of 3.2/11.8=27%. PCA found that two principal components accounted for 71% of the correlation structure among PAHs in moss, each associated with PAHs typically occurring in the particle and gas phase in the air, respectively. Based on correlations/loading with the principal components, we selected benzo(a)pyrene, pyrene, and naphthalene to further investigate associations with PAH-DNA blood adducts in linear regressions.

The absolute levels of PAH in moss do not have any particular meaning due to absence of reference ranges, but it is noteworthy that two different PAHs representative of gas- and particle-phases were detectable in all samples (namely: naphthalene and pyrene, respectively). The more volatile gas-phase naphthalene was more variable (CV of 16.3/13.1=124%) than the less volatile solid-phase pyrene (CV 10.1/28.5=35%). Benzo(a)pyrene, an established carcinogen, was detected in 42/53=79% of measurements. There was little evidence of either rank or linear correlation between any of the individual PAHs in moss and PAH-DNA adducts (details not shown).

**Table:** Descriptive statistics of measurements the levels of polycyclic aromatic hydrocarbons (PAH) in moss and blood among measurements above limit of detection; 53 samples collected for each measurement.

| Measurement | Number detected | Mean | Standard Deviation | Median | Minimum | Maximum |
| --- | --- | --- | --- | --- | --- | --- |
| *PAH blood adduct (/10<sup>8</sup>nucleotides)* | 32 | 11.8 | 3.2 | 11.5 | 7.9 | 22.9 |
| *PAH in moss (µg/kg)* |  |  |  |  |  |  |
| 1-Methylnaphthalene | 47 | 6.0 | 8.3 | 3.9 | 1.48 | 46.9 |
| 2-Methylnaphthalene | 53 | 6.6 | 9.5 | 5.3 | 2.32 | 72.4 |
| Acenaphthene | 28 | 4.1 | 4.0 | 3.2 | 1.77 | 23.5 |
| Acenaphthylene | 25 | 4.2 | 4.2 | 3.0 | 1.7 | 23 |
| Anthracene | 49 | 3.9 | 1.5 | 3.7 | 1.67 | 10.3 |
| Benzo(a)anthracene | 53 | 20.0 | 17.0 | 16.3 | 6.29 | 124 |
| Benzo(a)pyrene | 42 | 15.0 | 6.7 | 15.1 | 3.82 | 31.3 |
| Benzo(b)fluoranthene | 53 | 22.6 | 10.7 | 21.3 | 3.33 | 66.5 |
| Benzo(g,h,i)perylene | 49 | 22.7 | 9.8 | 21.1 | 7.625 | 68.5 |
| Benzo(k)fluoranthene | 52 | 15.9 | 5.3 | 15.3 | 6.57 | 29.6 |
| Chrysene | 53 | 38.6 | 12.8 | 36.1 | 14.2 | 65.6 |
| Dibenzo(a,h)anthracene | 14 | 10.4 | 4.8 | 9.1 | 4.525 | 20.6 |
| Dibenzofuran | 48 | 6.0 | 7.0 | 4.5 | 2.37 | 41.8 |
| Fluoranthene | 53 | 34.4 | 11.1 | 32.7 | 13.4 | 66.7 |
| Fluorene | 19 | 4.8 | 5.3 | 2.9 | 1.8 | 20.5 |
| Ideno(1,2,3-cd)pyrene | 35 | 11.0 | 4.7 | 9.9 | 3.23 | 25.7 |
| Naphthalene | 53 | 13.1 | 16.3 | 9.8 | 3.89 | 97.7 |
| Perylene | 49 | 16.6 | 6.3 | 16.2 | 2.43 | 39.5 |
| Phenanthrene | 53 | 20.9 | 5.7 | 21.4 | 10.1 | 35.6 |
| Pyrene | 53 | 28.5 | 10.1 | 29.1 | 10.75 | 68.4 |

Linear regression models with logarithm of PAH-DNA adduct as the dependent variable did not reveal any associations with PAH in moss. The unadjusted associations are illustrated in the **Figure**. Adjustment for age, race, gender, wood fires or dietary history did not substantially alter conclusion drawn from the **Figure**. The regression model containing all covariates and all three PAHs in moss had R^2^=0.2 with no indication of violation of the assumption of the analyses in the residual plots (not shown). The only notable association was consistent excess of PAH-DNA adducts in relation to wood fires in the house in preceding 48 hours, by a factor of about two (p=0.1). However, this was based on only three observations: in presence of wood fires, average level of PAH-DNA adduct was 12.9 *vs*. 8.0 adducts/10^8^nucleotided in absence of reported fires.

**Figure:**
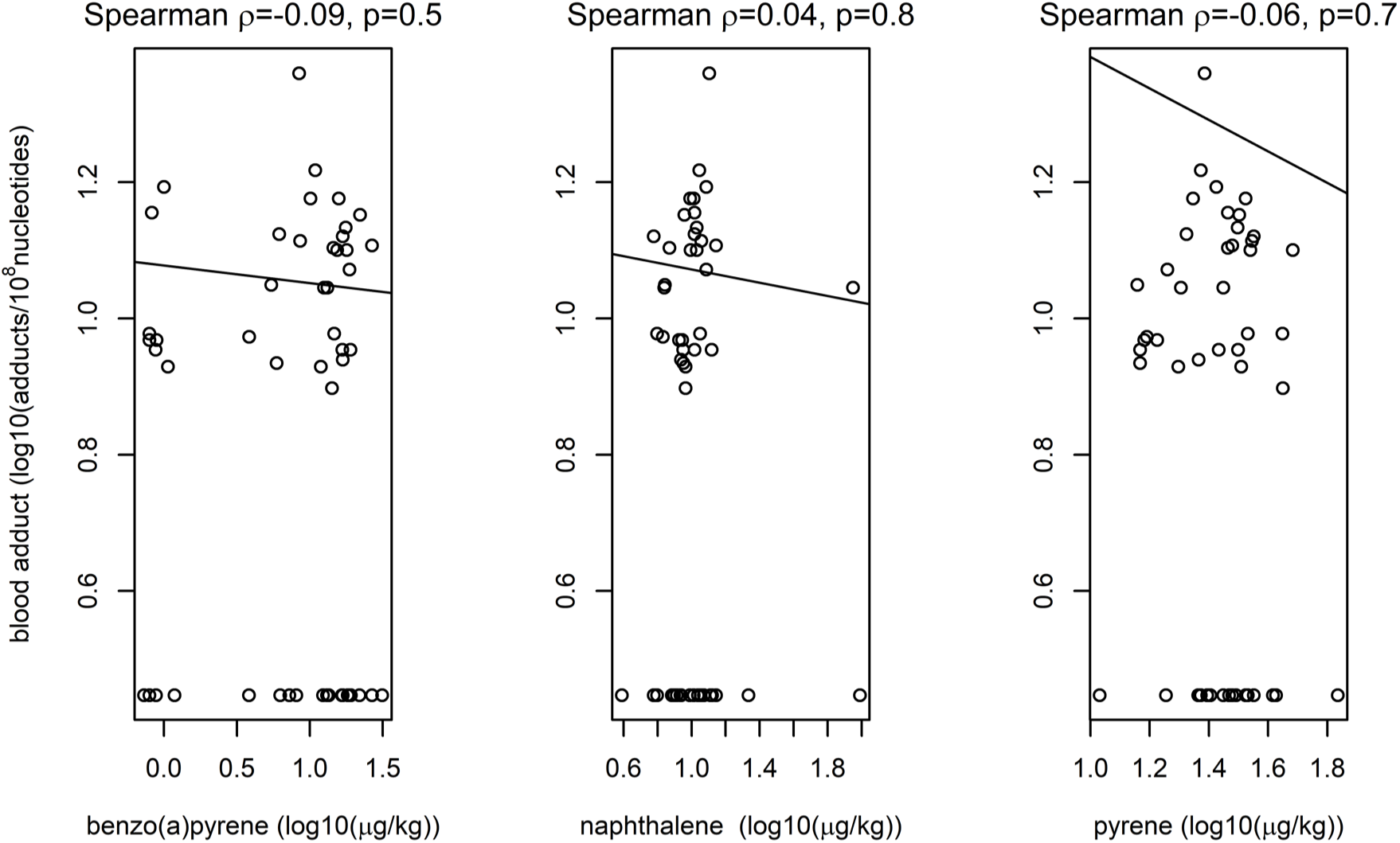
Linear models fit to logarithms of PAHs in moss and their PAH-DNA blood adducts along with rank correlations; non-detects of adducts and benzo(a) pyrene were imputed as half of the limit of detection.

## Discussion

We did not observe evidence to support the hypothesis that levels of PAH in the moss *O. lyellii* were related to levels of these exposures among nearby residents. Among the strengths of our approach was selection of non-smoking volunteers who resided with non-smokers, thereby controlling for a major source of confounding by design. That PAH-DNA adducts were potentially sensitive to at least one known source of PAH (wood fires) suggests these biomarkers may be useful for identifying more highly exposed individuals (11, 12). However, overall, we did not find that PAH levels in moss helpful in human exposure assessment in the studied setting.

Part of the reason for poor correlation of PAHs in moss and their adducts in human blood may be that the moss PAH concentrations were greatly influenced by sampling conditions (e.g., daily humidity and type of tree, local traffic conditions), whereas PAH-DNA adduct in human blood are subject to “physiological dampening” (13) that is less sensitive to these external conditions that can change daily (even if they are relevant). This is supported by our finding that the CV of adducts was lower than those of PAH measurements in moss. We attempted to account for some of these sources of confounding by limiting sampling to only one month and but we were not able to control for factors that Jovan et al. (7) saw as important to variation of PAH content of moss in the area. The levels of PAH in moss collected in May 2013 that we observed in the current study are substantially lower than those obtained using identical methods in Portland in December 2013 by Jovan et al. (7), underscoring the importance of seasonal trends and spatial variability.

Our study had several limitations. This study required relatively large moss samples (median cleaned and dried sample weight: 9.1 grams). While this was not a major challenge in Portland’s temperate climate, collecting samples of this size in other areas could be challenging. More sensitive laboratory techniques may overcome this limitation. One of the major limitations that we are not able to overcome with our data relates to differences in half-lives of PAH in moss and human PAH-DNA adducts. While PAH-DNA adducts have half-lives of about 3-4 months(14), the half-lives of PAHs in moss are not known. One study of temporal variability related particle-bound PAHs (such as benzo(a) pyrene and pyrene) in lichens to levels in the air two months prior(3) although due to their lower lipid content, moss is expected to represent shorter timeframes. If the two reservoirs of PAH (DNA adducts and moss content) integrate these levels over different timeframes, then a correlation would not be expected. This remains an important area of research that may help calibrate moss measurements to biomarkers in humans.

## Conclusion

Our research does not support the use of moss bio-indicators to assess risk of human exposure to PAH in Portland, Oregon.

## Data Availability

Data is not available.

## Acknowledgements

Thanks to Dr. Todd Rosentiel and Dr. Kenneth Stedman, Portland State University, for generously providing lab space. Consultation on PAH methods and extraction services provided by Specialty Analytical.

